# Decreasing Urinary Catheterization in Kidney Injury (DUCKI): an effectiveness and deimplementation study in the Intensive Care Unit

**DOI:** 10.1101/2025.01.19.25320779

**Authors:** Calvin Patten, Tiffany E Rosenzweig, Carrie Sona, AC Castro, Kristin Schmid, Maura Walsh, Laurie Robertson, Marilyn Schallom, Donna Prentice, Brian T Wessman, Enyo A Ablordeppey

**Author notes:** Enyo Ablordeppey MD MPH Washington University in St. Louis School of Medicine 660 South Euclid, Box 8054 St Louis, MO 63110. This study was performed at Washington University in St. Louis School of Medicine and Barnes-Jewish Hospital in St. Louis, MO.

## Abstract

**Funding:** EAA is funded by the Washington University Department of Anesthesiology’s Division of Clinical and Translational Research (DoCTR). Research reported in this publication was supported by the Washington University Institute of Clinical and Translational Sciences grant UL1TR002345 from the National Center for Advancing Translational Sciences (NCATS) of the National Institutes of Health (1). The content is solely the responsibility of the authors and does not necessarily represent the official view of the NIH.

**KEY POINTS:** This hybrid 1 implementation science study investigated the effectiveness of a program to reduce IUC utilization in ICU patients with AKI and ESRD. The DUCKI protocol successfully decreased IUC rates by 67% in the study’s targeted group of AKI and ESRD patients within an academic surgical ICU and was maintained over 2 years of follow up.

- Using implementation science to introduce evidence-based strategies like DUCKI is effective at increasing adoption and sustaining the practice.
- Oliguric patients offer a path of less resistance in changing catheterization practices.
- DUCKI can safely minimize IUC use in specific ICU populations.
- The protocol offers the potential for broader interventions to reduce catheter-associated risks in all ICU patients.

**BACKGROUND:** Indwelling urinary catheter use remains high in the surgical intensive care unit despite targeted, national efforts. When hospital-based initiatives occur, it is unclear if decreases in utilization are sustained.

**OBJECTIVES:** In 2021, we used implementation science to develop the Decreasing Urinary Catheters in Kidney Injury (DUCKI) program, targeting decreased indwelling catheterization in patients with acute kidney injury (AKI) with oliguria or end stage renal disease (ESRD). Three years later, we evaluated the effectiveness of DUCKI.

**METHODS:** This was a hybrid 1 implementation study. Outcomes of DUCKI eligible patients were evaluated through chart review with comparisons made between the 2021 and 2023 cohorts. Physicians and nurses were surveyed on the implementation effort.

**RESULTS:** ∼12.5% of patients were eligible for DUCKI. 70 patients in 6 months in 2021 and 19 patients in month in 2023 met DUCKI criteria. The average indwelling catheterization rate in DUCKI patients dropped to 10% from 80% in 2021. In 2023, the catheterization rate in DUCKI patients remains low (9%). Overall rates in the unit declined from 74% pre-implementation to 70% in 2021 and 66% in 2023. There were no serious adverse events associated with the protocol. The acceptability survey was completed by ICU stakeholders pre (n=88) and post (n=77) intervention. Respondents generally rated DUCKI positively, although a minority (26%) reported increased burden to workflow.

**CONCLUSIONS:** Low indwelling catheterization rates in patients with oliguric AKI or ESRD were sustained in the ICU’s DUCKI implementation program. This program has contributed to sustained decrease in overall unit catheterization.

## INTRODUCTION

Indwelling urinary catheter (IUC) utilization is one of the most performed in-hospital procedures, with reported daily usage rates of over 60% in Intensive Care Unit (ICU) patients (2, 3). Common indications for IUC include urinary retention, need for close monitoring of urinary output, and perioperative use (4, 5). However, significant portions of patients who do not meet such criteria have previously been identified, with studies indicating that 38-46% of IUC-days have no appropriate indication, including in ICUs (6–8). IUC overuse is associated with adverse events (for example, pain, hematuria, delirium) and complications such as Catheter Associated Urinary Tract Infections (CAUTI)(3, 5, 9–11). Most hospital acquired UTIs are catheter associated, 65% of which are preventable, and contribute to excess mortality and financial impact (12–16). Despite fiscal motivation for hospital systems to decrease IUCs and CAUTIs, there is limited success based on the Centers for Medicare and Medicaid Services (CMS) payment withholding strategy (15–17). Similarly, attempts to decrease IUC utilization rates in the ICU have failed or been unsustained, despite targeted, best practice implementation efforts (2, 18, 19). Reasons for this are unclear and likely multifactorial.

One potential area of IUC overuse is in the management of patients with oliguric acute kidney injury (AKI) requiring renal replacement therapy or end stage renal disease (ESRD). Incidence varies depending on location and ICU specialization, but prior work has shown that 13-17% of patients undergo renal replacement therapy (RRT) for AKI during their first week of ICU admission, while 3-14% of patients admitted to the ICU have pre-existing RRT dependence because of ESRD (20–22). IUCs may be placed and maintained in these patients (up to 30% ICU patients) reflexively without indication, or specifically to monitor potential renal recovery. However, effective and less invasive options to monitor renal recovery exist, including non-invasive Bladder Scanner® or 2D ultrasound (23, 24). We identified this population of patients with ESRD and AKI as a potentially high yield, well-defined group, to target our implementation initiative that might inform more generalized strategies to reduce unnecessary catheterization across all patients in the ICU. The goal of this project was to use an implementation science approach to change the behavior of clinicians to decrease IUC use in patients with oliguric AKI and ESRD.

## METHODS

We took a hybrid 1 implementation science approach to evaluate the effectiveness of the intervention and the outcomes of implementation (25, 26). Hybrid 1 studies prioritize effectiveness but still collect implementation outcomes like acceptability. We selected implementation strategies that were informed by barriers using the consolidated framework for implementation research (CFIR) and used Proctor’s taxonomy of implementation outcomes to guide the selection of outcomes and measurements (26–28). We aimed to understand the effectiveness of an implementation program targeting IUC removal in the high yield patient group through a novel designed program called DUCKI (Decreasing Urinary Catheters in Kidney Injury). We also studied the unit’s acceptance of the protocol via survey pre- and post-implementation to improve program adoption and sustainability.

Our group initiated the DUCKI quality improvement program in a 36-bed surgical intensive care unit (SICU) in an urban, academic medical center. This study is a hybrid type 1 study of a SICU implementation program that prevents catheterization in ESRD patients and/or promotes timely removal of IUC as illustrated in **Figure 1**. This project was designed to comply with quality standards for survey reporting in medical literature. Review of data collected as part of routine clinical practice in this study were in accordance with the ethical standards of the Helsinki Declaration of 1975 and reviewed by the Institutional Review Board of Washington University.

**Figure 1.**
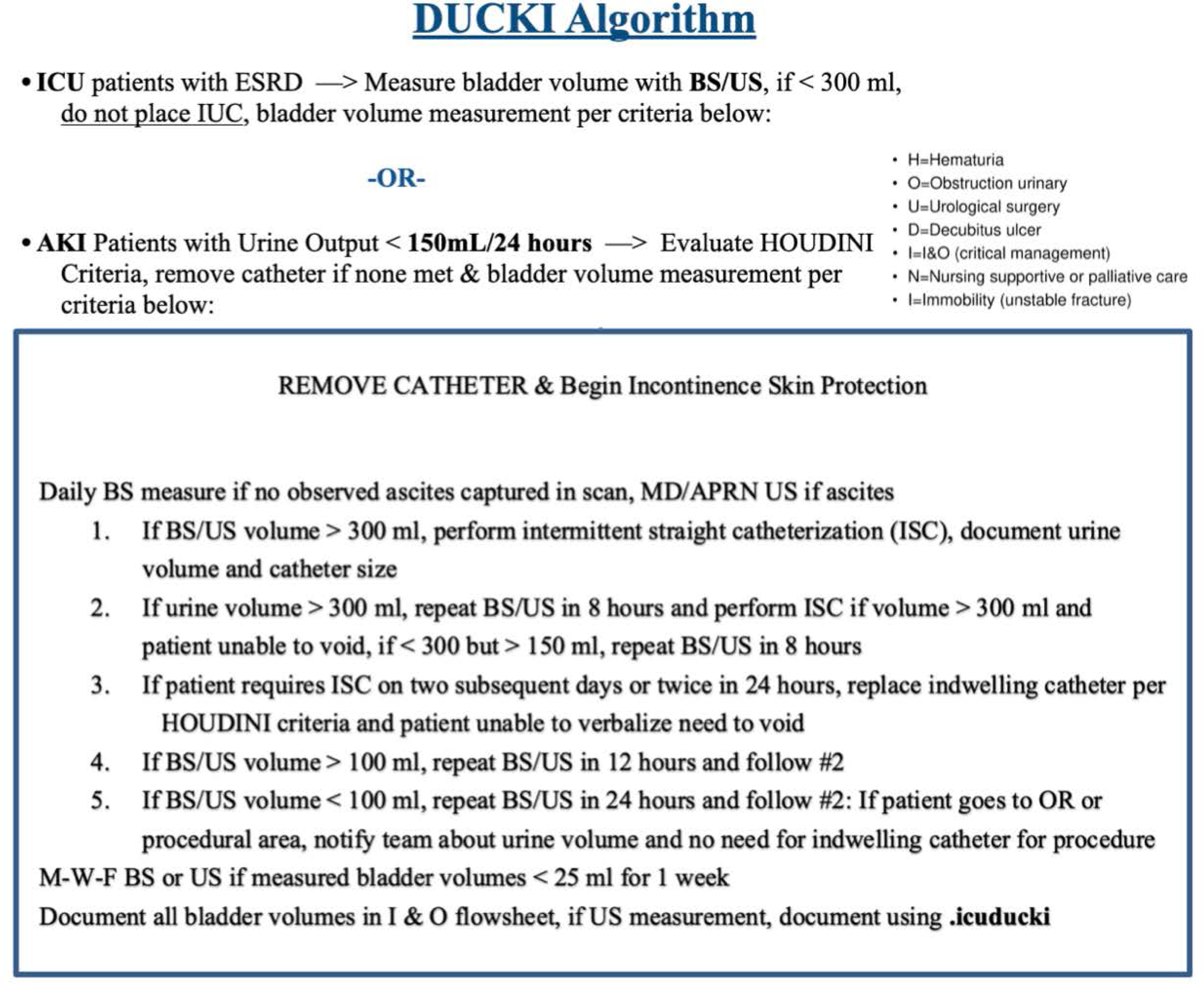
DUCKI algorithm to decrease indwelling urinary catheters in kidney injury patients. DUCKI, Decreasing Urinary Catheter in Kidney Injury; ICU, intensive care unit; ESRD, end stage renal disease; BS, bladder scanner; US, 2-D ultrasound; IUC, indwelling urinary catheter; AKI, acute kidney injury; MD, medical doctor; APRN, advance practice registered nurse (or equivalent); OR, operating room

### DUCKI Algorithm

The research team designed DUCKI to supplement the HOUDINI protocol, a nurse-led practice in removing IUCs within an acute healthcare setting based on criteria, to the team decision-making structure of the hospital [8,10,12]. Despite low urine output among ESRD and oliguric AKI patients, the catheterization rate among this group was 80% with catheters remaining in place just as they would for other patients. The research team observed that physicians were reluctant to lose the output data point, even if the number was always null. Additionally, some RNs reported preferring IUC to external catheters for those with urine output. Through conversations with stakeholders (ICU leadership, ICU quality improvement committee, nephrology, and urology), the algorithm in **Figure 1** was developed, and the program was fully approved by stakeholders and initiated in March 2021. Eligible patients included ESRD patients with <300ml of urine in 24 hours or AKI patients with < 150 ml of urine in 24 hours (oliguria). DUCKI program patients would not have an IUC placed upon ICU admission. Those with existing IUCs could have catheter automatically removed by nursing barring exclusions from the *HOUDINI* catheter removal criteria, surgical procedure via laparoscope, or surgical team indication for IUC within the first two postoperative days (29). After removal, bladder volume was monitored by a Bladder Scanner® or 2D ultrasound a minimum of once per 24 hours. Bladder scanning has been demonstrated to be accurate for estimating urine volume, identifying retention, and reducing the need for unnecessary catheterization (30–32). For greater accuracy, patients with ascites were required to receive 2D Ultrasound monitoring (24).

### Outcome Measures

Outcomes in the first 6 months included IUC insertion and removal dates, incidence of IUC reinsertion, and return urine output in AKI patients as measured by the research team. We calculated a IUC utilization rate for eligible patients. We also tracked potential adverse events such as bladder rupture, moisture associated skin damage, or obstructive uropathy. At the ICU level of analysis, unit utilization rate and CAUTI rate was collected 5 months prior to DUCKI and monthly during implementation as reported by the unit’s quality improvement (QI) committee.

We identified the protocol’s acceptability to the ICU team as a driver in sustainability and therefore a measure of success (33). To evaluate this, we designed an electronic survey based on stakeholder (ICU medical director, nursing leadership, critical care faculty physician) feedback. While no existing acceptability instruments addressed urinary catheters or low-value health practices specifically, the questions were guided by CFIR and the outcome definitions established by Proctor et al. After pilot testing to avoid bias, it was issued to critical care medicine physicians and nursing for voluntary completion pre-implementation and at 3 months post-implementation. Using a Research Electronic Data Capture (REDCap®), the survey assessed attitudes and barriers to IUC removal, as well as acceptability of the protocol. Data was regularly reported back to the stakeholders.

### DUCKI Implementation

Selected implementation strategies for this program (**Figure 2**) were informed by the barriers identified in previous qualitative work using CFIR, primarily concerns about pushback from outside the unit, failure of external catheters especially for female patients, inaccurate measurement of urine output, and lack of time in addition to lack of knowledge about bladder scanning and the specific patient populations. The strategies implemented were 1) education and training on the algorithm and bladder volume measurement by Bladder Scanner® and/or 2D Ultrasound, 2) decision support to ICU teams, 3) audit and feedback to the ICU QI committee, 4) organizational support (policy/procedure) from urology and nephrology subspecialities as well as ICU leadership, 5) diverse implementation team, and 6) algorithm development and dissemination (34). The DUCKI implementation team provided both in-person and online training on the IUC removal algorithm to ICU doctors and RNs. The program was regularly discussed during unit meetings.

**Figure 2.**
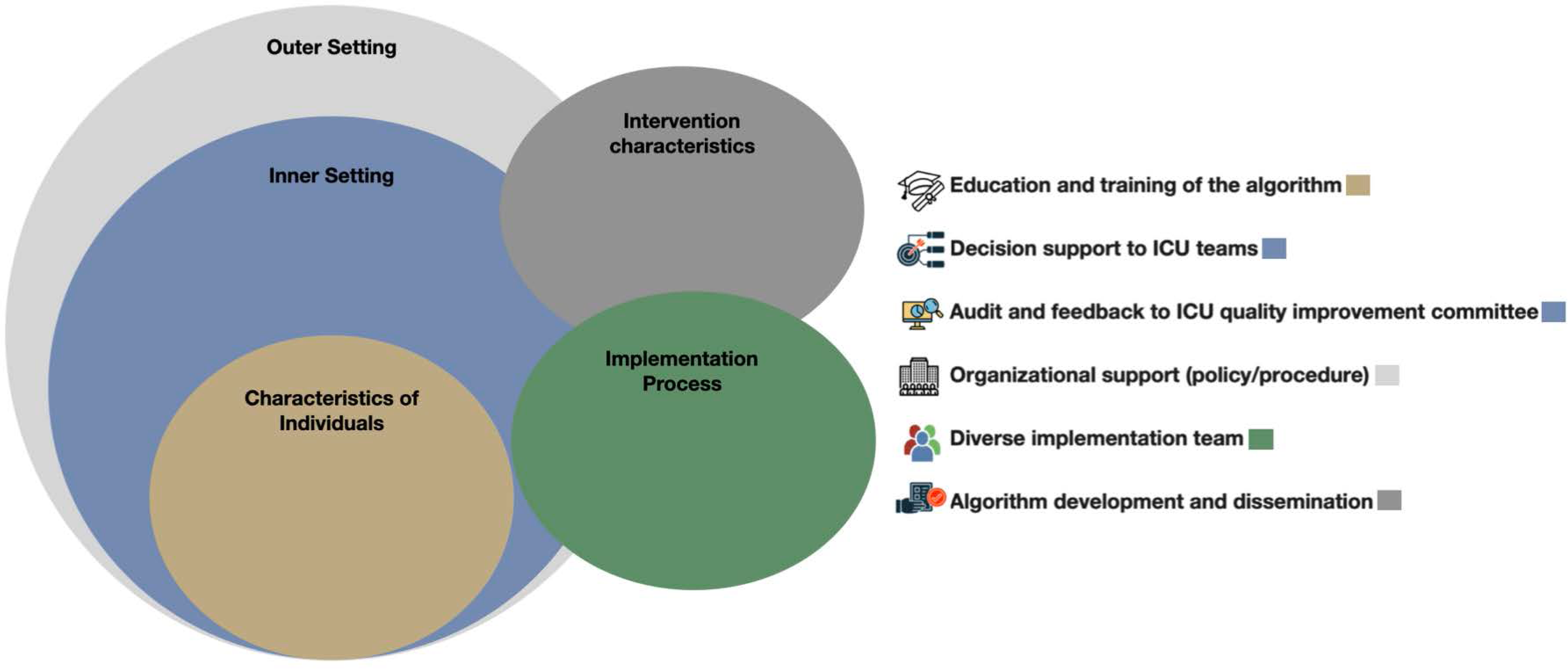
Six selected implementation strategies for DUCKI program. CFIR is used to identify barriers to reducing IUCs through DUCKI program. Each strategy is mapped to appropriate CFIR domain level (circles). ICU, intensive care unit; DUCKI, decreasing urinary catheter in kidney injury; CFIR, consolidated framework in implementation research; IUC, indwelling urinary catheter

Over the initial 6 months of implementation, research team nurses (MS, DP, CS) reviewed the daily patient census, identified patients who met the DUCKI criteria, assessed their outcomes, and monitored clinicians’ adherence to the protocol. In addition to chart review, the research nurses (CS, LR) physically confirmed the presence of dialysis machines and talked with the RNs on duty about patients who were eligible. RNs initiated conversations about IUC necessity during ICU team rounding discussions using the daily ICU rounding checklist to drive protocol adherence. Members of the research team performed this review intermittently after the 6-month daily chart review ended. A full review of all SICU patients was conducted 3 years later for 4-weeks (July 12 – August 7, 2023) using the same process in 2021 to measure the same outcomes and determine DUCKI program sustainability (33).

## RESULTS

### Chart Review

There were 70 patients over 6 months in 2021 and 19 patients in one month during 2023 who met the criteria for DUCKI, representing approximately 9% of the 36 bed ICU population each day. Demographic and unit data is reported in **Table 1** showing 36 patients (51%) and 13 (68%) had ESRD while 34 (49%) and 6 (32%) had AKI in 2021 and 2023 respectively. In Table 2, overall IUC rates in the SICU declined from 74% in 2020 pre-implementation to 70% in 2021 and now 66% in 2023. This 8% reduction in overall IUC rate (9% DUCKI eligible x 12% IUC) is attributed to decreased rates in DUCKI patients. CAUTI rate/1000 catheter days declined from 1.5 in 2021 to 0.8 in 2023. Given the low frequency of CAUTI and the array of factors that contribute to their occurrence, it is not possible to infer details about changes in CAUTI rates from this data cohort.

**Table 1.** Patient Demographics and Outcomes.

|  | 2021 (%) | 2023 (%) | Total (%) |
| --- | --- | --- | --- |
| Total Eligible Patients | 70 (79) | 19 (21) | 89 (100) |
| Age (years) | 58.7 | 62.4 | 60 |
| Male n(%) | 44 (63) | 12 (63) | 56 (63) |
| Female n(%) | 26 (37) | 7 (37) | 33 (37) |
| ESRD n(%) | 36 (51) | 13 (68) | 49 (55) |
| Catheter Replaced | 1 (3) | 0 (0) | 1 (2) |
| Required ISC | 3 (8) | NA | 3 (5) |
| AKI w/ UOP <150mL/24 hour n(%) | 34 (49) | 6 (32) | 40 (45) |
| Renal Recovery | 12 (35) | 3 (50) | 15 (38) |
| Catheter Replaced | 7 (21) | 1 (17) | 8 (20) |
| Required ISC | 13 (38) | NA |  |
| Patient Complications | 70 (79) | 19 (21) | 89 (100) |
| MASD | 3 (4) | 0 (0) | 3 (3) |
| Obstructive Uropathy | 0 (0) | 1 (5) | 1 (1) |
| Bladder Rupture | 0 (0) | 0 (0) | 0 (0) |

Data from 2021 incorporates 6 months; data from 2023 incorporates 1 month of chart review.
ESRD- end stage renal disease; ISC- intermittent straight catheterization; AKI- acute kidney
injury; UOP- urinary output; MASD- moisture associated skin damage

**Table 2.** DUCKI program outcomes.

| DUCKI Program Application | 2020 | 2021 | 2023 |
| --- | --- | --- | --- |
| ESRD: IUC Removed (%) | n/a | 25 (70) | 6 (46) |
| ESRD: IUC Never Placed (%) | n/a | 11 (30) | 7 (54) |
| Unit IUC Utilization Rate | 74% | 70% | 66% |
| CAUTI Rate per 1000 Cath Days | 1 | 1.5 | 1.1 |
| Daily Catheterization of DUCKI<br>Eligible Patients | 80.0% | 12.5% | 8.7% |

Data from 2021 incorporates 6 months; data from 2023 incorporates 1 month of chart review.
DUCKI-decreasing urinary catheter in kidney injury program; ESRD- end stage renal disease;
IUC-indwelling urinary catheter; CAUTI-catheter associated urinary tract infection

DUCKI patient outcomes are reported in **Table 3**. Among the AKI patients, 12 (36%) in 2021 and 3 (50%) in 2023 experienced a return of kidney function with high urine output. Of the patients with AKI, 8 of the 15 had an IUC replaced after return of kidney function; the other patients did not require the use of a catheter, indwelling or external. The average daily catheterization rate in the AKI/ESRD patient group pre-DUCKI was 80% and decreased to 12.5% during 6-month implementation. In a sample of the 2023 SICU chart review, we found a catheterization rate of 8.7% in the AKI/ESRD group. There were no serious adverse events noted among the patients who participated in DUCKI. In 2021, 16 patients required intermittent straight catheterization (ISC) for return of urine output (18.6%) and inability to void (4.3%). IUCs were replaced in 8 AKI patients (11.4%) following returned urine output after 2 uses of ISC. One ESRD patient requested an IUC be reinserted in lieu of using ISC every few days. Incontinence was noted in 10 patients (14.3%), and two of those patients developed dermatitis or moisture associated skin damage (MASD). An incidental case of hydronephrosis was identified on CT imaging in 1 patient.

**Table 3.** Acceptability Survey Question (pre/post)

| DUCKI program component perception |  | Pre-implementation Agreement (%) | Post-Implementation Agreement (%) |
| --- | --- | --- | --- |
|  | Total | 88 | 74 |
|  | MD | 15 | 8 |
|  | RN | 73 | 66 |
| Implementation of a urinary catheter removal protocol in patients with acute kidney injury and end stage renal disease who produce minimal urine (would be/is) beneficial to patients | Total | 79 (90) | 69 (93) |
|  | MD | 15 (100) | 8 (100) |
|  | RN | 64 (88) | 61 (92) |
| By following the urinary catheter removal protocol, I have the ability to decrease urinary catheter days in the SICU patient population | Total | 83 (94) | 71 (96) |
|  | MD | 15 (100) | 8 (100) |
|  | RN | 68 (93) | 63 (96) |
| I have the ability to decrease Catheter Associated Urinary Tract Infections (CAUTI) in the SICU patient population. | Total | 81 (92) | 71 (96) |
|  | MD | 13 (87) | 8 (100) |
|  | RN | 68 (93) | 63 (96) |
| I am confident I can accurately measure urinary bladder volume with a bladder scanner or point of care ultrasound. | Total | 70 (80) | 59 (79) |
|  | MD | 12 (80) | 3 (38) |
|  | RN | 58 (79) | 56 (85) |
| I consider the addition of a urinary catheter removal protocol a burden to my work schedule | Total | 15 (17) | 19 (26) |
|  | MD | 6 (40) | 0 (0) |
|  | RN | 9 (12) | 19 (29) |
| I (would incorporate a/incorporate the DUCKI) urinary catheter removal protocol in my daily practice to reduce CAUTI in the SICU | Total | 81 (92) | 58 (78) |
|  | MD | 13 (87) | 7 (88) |
|  | RN | 68 (93) | 51 (77) |
| Rate your current knowledge of (the/the DUCKI) urinary catheter removal protocol in the SICU that includes Acute Kidney Injury patients:0=No Knowledge -10 | Total | 4.8 | 6.7 |
|  | MD | 3.1 | 5.75 |
|  | RN | 5.2 | 6.81 |
| Please indicate how many times per week you use the bladder ultrasound or point of care ultrasound to measure a patient's bladder volume (MD Only) | Never | 9 (60) | 5 (63) |
|  | 1-5 | 5 (33) | 3 (38) |
|  | 6-10 | 1 (7) | 0 (0) |
|  | >10 | 0 (0) | 0 (0) |
| Please indicate how many times per week you use the bladder scanner to measure a patient's bladder volume (RN Only) | Never | 11 (15) | 6 (9) |
|  | 1-5 | 59 (81) | 58 (88) |
|  | 6-10 | 2 (3) | 2 (3) |
|  | >10 | 1 (1) | 0 (0) |

MD- medical doctor; RN- registered nurse; DUCKI-decreasing urinary catheter in kidney injury program; SICU- surgical intensive care unit

### Survey

The total response rate of 131 ICU stakeholders surveyed was 67% pre and 56% post. Eighty-nine percent of participants were RN’s. Seventy-six percent of nurses and 88% of physicians reported daily participation in the program since implementation. In both pre and post implementation surveys (Table 4), over 93% of RN and 100% of physician respondents indicated that following a program to remove catheters in patients with ESRD and oliguric AKI would be beneficial for patients, with similarly high percentages anticipating that they could reduce the occurrence of CAUTI and decrease IUC days. Those citing an IUC removal program as a burden increased from 12% to 29% of RNs post-implementation, while physicians fell from 40% to zero percent. 77% of RNs and 88% of physicians reported using DUCKI daily. In the post-implementation survey, the most cited barriers to DUCKI were concern for skin breakdown in female patients (72%), inaccurate urine output measurement (57%), and resistance to catheter removal by the surgical (49%) or ICU (39%) teams.

## DISCUSSION

In this select group of patients with oliguria and either preexisting ESRD or AKI, our implementation program successfully and dramatically reduced IUC usage in an academic surgical ICU without adverse events. Furthermore, follow up data collection 2 years later supports that the program has been sustainable. We believe this is the first study to focus exclusively on IUC minimization on this subgroup of ICU patients using implementation science strategies and outcomes.

First, this study demonstrates the successful use of an implementation science approach with qualitative interviews combined with targeted strategy selection to change clinical behavior. Prior interventions to reduce IUC use have been inconsistent in adaptation or quickly waned after the active implementation period (2, 13, 18, 19, 35–39). These interventions often focused on educational interventions but may not have adequately addressed structural and individual characteristics, which encompassed the most identified barriers (40). Menegueti et al demonstrated at education plus feedback was necessary to see immediate and sustained decrease in IUC use(41). We used behavioral theories and principles to select appropriate strategies that mitigate barriers and leverage facilitators (25, 26, 28, 33). We addressed structural barriers by ensuring the availability of bladder scanners and identifying a population that would not require significant additional time(42). The result was a greater than 60% reduction of IUC use in the target population that contributed to an 8% decline in IUC use, which persisted 3 years later.

Similarly, the pre-implementation survey responses suggested that providers found a high level of acceptability and appropriateness with the algorithm, which we leveraged in implementation. A 6-month follow-up survey demonstrated a high rate of self-reported adoption. The surveys functioned in part to engage stakeholders and identify barriers and facilitators to implementation, while also supporting dissemination. DUCKI included providing frequent education about IUC harms and alternatives and offering small incentives for staff who removed IUCs. The initial implementation effort included the use of observation to ensure fidelity and compliance with the algorithm, which supported the tailored strategy of audit and feedback. While intermittent quality checks and onboarding education persist, the use of incentives, observation, and audit and feedback have been largely discontinued. Our 2023 screening demonstrated similar IUC utilization to 2021, suggesting that sustainability may be driven by changes in unit culture and work infrastructure. Indeed, despite a decrease in implementation efforts after 6-12 months, IUC necessity remains a topic of rounds.

Second, changes in behavior by clinical providers to avoid or remove IUCs were not associated with severe adverse events, but minor events were observed. Two cases of MASD in female patients were identified via chart review, potentially reflective of staff concerns regarding external collecting devices for women. Additionally, an AKI patient receiving an unrelated computed tomography scan was identified as having bilateral hydronephrosis with 650cc of retained urine. On review of this patient’s records, this was likely related to inconsistent bladder scanning and non-adherence to the DUCKI program. In the setting of IUC removal in AKI patients, return of renal function was consistently recognized with scheduled bladder scanning, and in the appropriate situation catheters were reinserted at that time. Notably, none of our patients experienced bladder rupture.

Finally, we simplified the decision process by clearly identifying a population that does not require an IUC. The predominantly RN group responded in both surveys that a dedicated program would help reduce IUC use and preventing CAUTI, highlighting motivation related to patient outcomes. In this RN group, 76% reported utilizing our DUCKI program daily, emphasizing engagement from bedside staff. Notably, a minority of our staff did report that the algorithm was a burden on their workflow, but at least a portion of this cohort used it daily despite these reported challenges. Future adaptation should focus on barriers such as concerns about skin breakdown or resistance beyond the ICU team.

## LIMITATIONS

Our study has several limitations. It was performed at a single academic surgical ICU, and its generalizability to other ICU environments has not been evaluated. While a decrease in catheterization days was observed compared to historical data, this is a retrospective assessment without a contemporary control group for comparison. We are unable to assess the impact of this algorithm on CAUTI rates given the relatively low frequency of CAUTI and limited sample size. Patients were not followed up with after discharge from the ICU, potentially underestimating the harms of IUC removal. Finally, this is a very specific group of patients, the vast majority of whom required dialysis, and not generalizable to all ICU patients with AKI. Future work will be required to evaluate the reproducibility of this implementation program in other ICUs and across other patient cohorts. Behavioral modification lessons from this study can inform further interventions for the broader ICU patient population to enhance appropriate IUC removal and minimize the potential harms of unnecessary catheterization in all ICU patients.

## CONCLUSIONS

Removal of IUCs in patients with AKI and avoidance of placement in ESRD can be performed safely with non-invasive bladder volume monitoring thus eliminating the risk of CAUTIs. Low IUC rates in patients with oliguric AKI or ESRD is well accepted and sustained in the overall SICU population with the DUCKI (IUC removal) implementation program. Implementation science helped formulate our DUCKI program and was paramount to its success and long-term sustainability. There are notable barriers to IUC removal that need further exploration to guide additional strategy development and be generalizable to IUC removal efforts for all ICU patients.

## Supporting information

Supplemental File 1, STROBE checklist

## Data Availability

All data produced in the present study are available upon reasonable request to the authors

